# Replication of an open-access deep learning system for screening mammography: Reduced performance mitigated by retraining on local data

**DOI:** 10.1101/2021.05.28.21257892

**Authors:** J.J.J. Condon, L. Oakden-Rayner, K.A. Hall, M. Reintals, A. Holmes, G. Carneiro, L.J. Palmer

## Abstract

**Aim:** To assess the generalisability of a deep learning (DL) system for screening mammography developed at New York University (NYU), USA (1, 2) in a South Australian (SA) dataset.

**Methods and Materials:** Clients with pathology-proven lesions (n=3,160) and age-matched controls (n=3,240) were selected from women screened at BreastScreen SA from January 2010 to December 2016 (n clients=207,691) and split into training, validation and test subsets (70%, 15%, 15% respectively). The primary outcome was area under the curve (AUC), in the SA Test Set 1 (SATS1), differentiating invasive breast cancer or ductal carcinoma in situ (n=469) from age-matched controls (n=490) and benign lesions (n=44). The NYU system was tested statically, after training without transfer learning (TL), after retraining with TL and without (NYU1) and with (NYU2) heatmaps.

**Results:** The static NYU1 model AUCs in the NYU test set (NYTS) and SATS1 were 83.0%(95%CI=82.4%-83.6%)(2) and 75.8%(95%CI=72.6%-78.8%), respectively. Static NYU2 AUCs in the NYTS and SATS1 were 88.6%(95%CI=88.3%-88.9%)(2) and 84.5%(95%CI=81.9%-86.8%), respectively. Training of NYU1 and NYU2 without TL achieved AUCs in the SATS1 of 65.8% (95%CI=62.2%-69.1%) and 85.9%(95%CI=83.5%-88.2%), respectively. Retraining of NYU1 and NYU2 with TL resulted in AUCs of 82.4%(95%CI=79.7-84.9%) and 86.3%(95%CI=84.0-88.5%) respectively.

**Conclusion:** We did not fully reproduce the reported performance of NYU on a local dataset; local retraining with TL approximated this level of performance. Optimising models for local clinical environments may improve performance. The generalisation of DL systems to new environments may be challenging.

**Key Contributions:** In this study, the original performance of deep learning models for screening mammography was reduced in an independent clinical population.

Deep learning (DL) systems for mammography require local testing and may benefit from local retraining.

An openly available DL system approximates human performance in an independent dataset.

There are multiple potential sources of reduced deep learning system performance when deployed to a new dataset and population.

## Aim

To assess the generalisability of a previously published open-access deep learning (DL) system for mammographic breast cancer screening, developed on a US clinical population at New York University (NYU)(1, 2), in the public, mammographic screening service of South Australia (SA).

## Background

Breast cancer is responsible for 1 in 4 cancer deaths in women worldwide.(3) Early detection and treatment of breast cancer, when the tumour is smaller and less likely to have metastasised, is associated with improved survival.(4) Delays in diagnosis and treatment are associated with adverse clinical outcomes.(5) Later detection and consequently more advanced cancers often require more intensive treatments that can decrease quality of life.(6) An Australian case-control study found a relative reduction in breast cancer mortality of 30-41% associated with screening participation.(7) Randomised controlled trials have demonstrated relative reductions of 20-30% in breast cancer mortality attributable to mammographic screening programs, for women aged 50-69 years (8–11) and in women from age 40.(12)

BreastScreen Australia is the national, public screening program for unsuspected breast cancer and has operated since 1991. It aims to reduce illness and death from breast cancer and to enable early intervention.(13) BreastScreen South Australia (BSSA) operates the public screening program, including mammography, in the state of South Australia. BSSA invites women resident in South Australia, primarily aged 50 to 74, to undergo screening mammography exams every two years at one of seven stationary and three mobile clinics. The diagnostic accuracy of human radiologists interpreting screening mammography has been reported in a seminal study as a partial AUC of 88%.(14)

Convolutional neural networks (CNNs) are a family of deep learning methods (15) which are especially suited to analyse complex data such as images. With labelled examples to learn from, these models are automatically optimised to improve performance at the given task (“training”).(15) Training optimisation occurs via updates to the numerical parameters (or weights) of a model.(15) Rather than starting training with random weights (random initialisation or training “from scratch”), weights can be transferred from another learnt task and retrained (“transfer learning”).(16)

Recent studies have reported human or above human-levels of performance with CNNs for screening mammography.(2, 17–20) However, one study has found that the performance of a commercial CNN system (21) was reduced and inferior to humans when tested in another country and population to that of its training.(22) Some authors have reported inconsistent performance of deep learning models on mammogram classification, with AUCs ranging from 88-95%.(23) CNN performance has been shown to be sensitive to dataset characteristics, image idiosyncrasies, and the distribution of the training data,(24–26) which may account for reduced performance in some situations. For example, there is evidence that the performance of CNNs can vary based on the vendor of the mammography equipment.(27) Some authors have suggested that adjusting CNNs with local data may improve performance.(17) The capacity of CNNs to maintain their performance on new and independent data, including from new locations, is referred to as ‘generalisability’. To investigate the limitations of DL generalisability in the context of screening mammography, we tested an openly available DL system developed by Wu *et al*. at NYU (1, 2) in the BSSA dataset.

## Methods and Materials

The current study was approved by the Central Adelaide Local Health Network Institutional Review Board (HREC/16/RAH/229, R20160601), with a waiver of consent for the retrospective use of deidentified clinical data. Women with biopsy and surgical pathology-proven lesions (n=3160) and age-matched controls (n=3240) were selected from all women screened at BSSA with full-field digital mammography from January 1 2010 to December 31 2016 (n clients=207,691, see Figure 1). Clients were randomly split into training, validation and test subsets (70%, 15%, 15% of the BSSA dataset respectively), stratified for the dominant finding of the dominant lesion, based on further lesion characterisation with mammographic and ultrasound imaging. See Table 1 and supplementary material for further dataset characteristics. The primary balanced test set (Test Set 1) included women with invasive breast cancer (IBC) or ductal carcinoma in situ (DCIS) (n=469), benign lesions (n=44) and age-matched controls (see Table 1, figure 1, Table S1 and S3). All case subsets were age-matched from a pool of control clients. Where there were multiple potential controls of the same age, the control was randomly selected. Clients with less than the four standard views - left and right craniocaudal (CC) and mediolateral oblique (MLO) - were removed from any training or analysis. We excluded any client with previous malignancy or biopsy, implants or breast symptoms from the control pool. Control clients with only one round of screening were excluded and only exams with at least one subsequent follow-up (where there were no concerning abnormalities) were included (see figure 1).

**Table 1.**
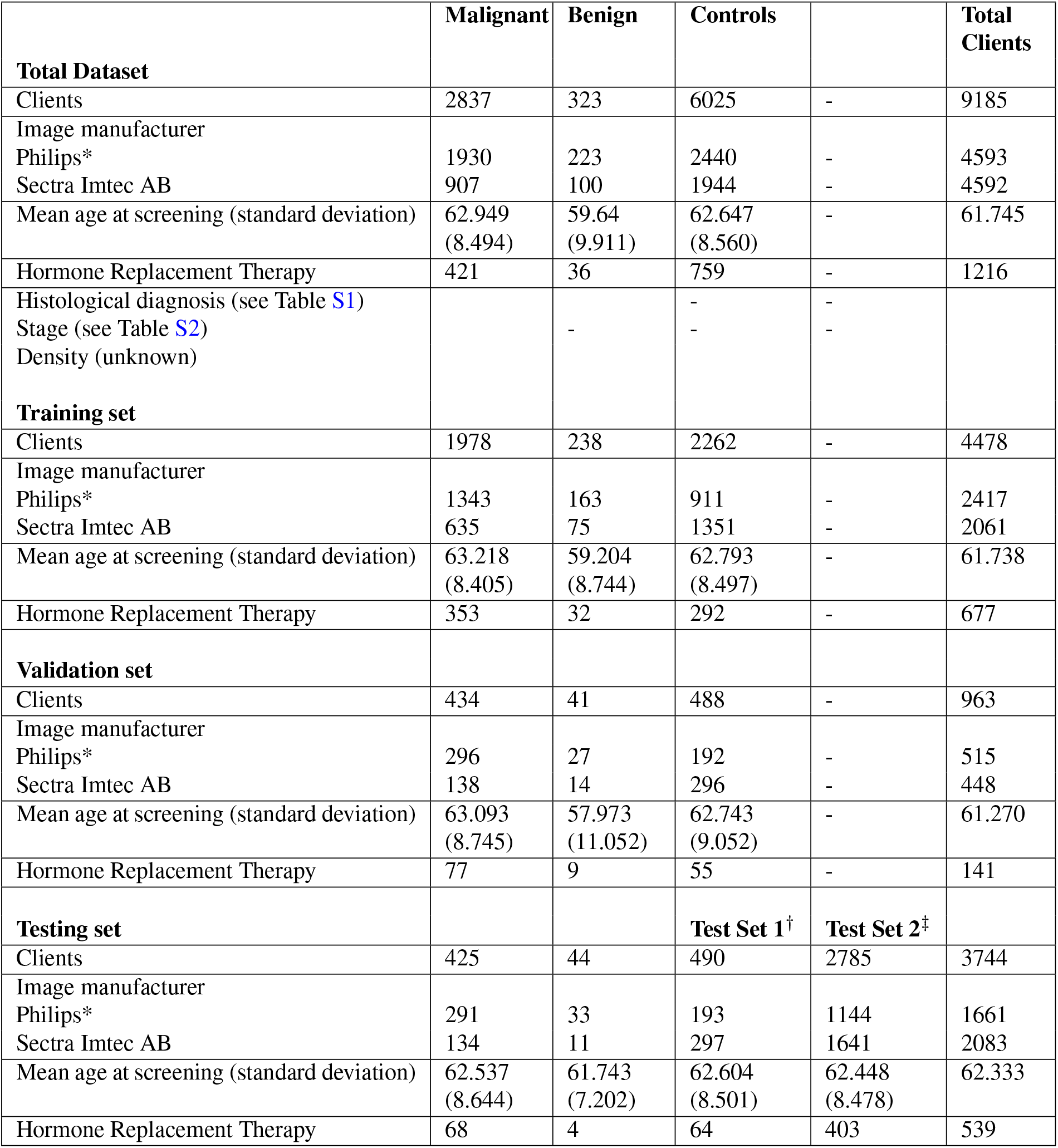
BSSA Dataset. *Philips Medical Systems and Philips Digital Mammography Sweden AB. †: Test Set 1 - balanced controls. ‡: Test Set 2 - approx. NYU prevalence controls.

**Fig. 1.**
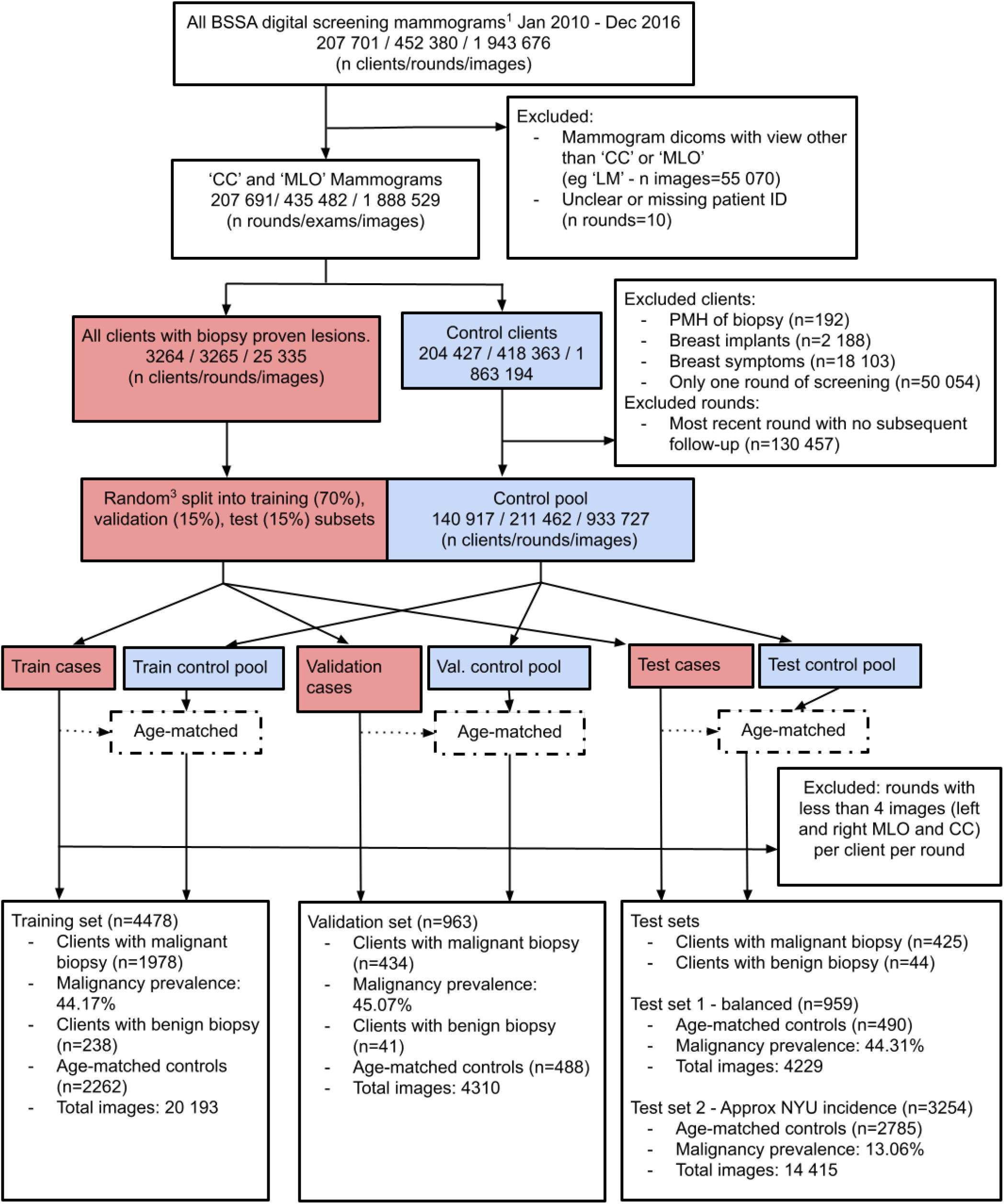
Dataset flow diagram. 1. Excluding assessment mammograms. 2. We use ‘round’ to refer to one episode of screening, consisting of at least four standard views (‘CC’ and ‘MLO’ for each breast). 3. Stratified client-wise by dominant finding.

To utilise the NYU models, all available biopsy-proven benign cases were included with their own label for model training and hyperparameter validation (see Figure 1). To match the clinical task of interpreting screening mammograms, the primary outcome of interest was differentiating IBC or DCIS from age-matched controls and biopsied, benign lesions. For all experiments, receiver operating characteristic (ROC) curves and the area under these curves (AUC) were calculated. Confidence intervals were calculated using the bootstrap method (28), resampling test clients with replacement for ten thousand iterations.

All modelling and analyses were performed on a single, physical workstation with 4TB of HDD, 2TB of NVME SSD, an Intel® Core™ i9-7920X CPU @ 2.90GHz × 24 with 62GB of RAM and 2 x GeForce RTX 2080TIs (12GB each).

### A. Replication of Static NYU Models

The model and preprocessing code were run as published (1) on Test Set 1, although some additional preprocessing was necessitated by differences in image acquisition, storage and size (see Supplementary Note 2 and Figures S4 and S5).

Patch-wise benign and malignant heatmaps were generated per image (see Figure 2) using a CNN trained on NYU radiologists’ hand drawn benign (n=3 158) and malignant (n=855) segmentations, produced by Wu *et al*. (2), which is freely available (1). Inference of this network calculates a probability of each patch belonging to benign and malignant segmentations. These probabilities were overlayed on original images (see Figure 2).

**Fig. 2.**
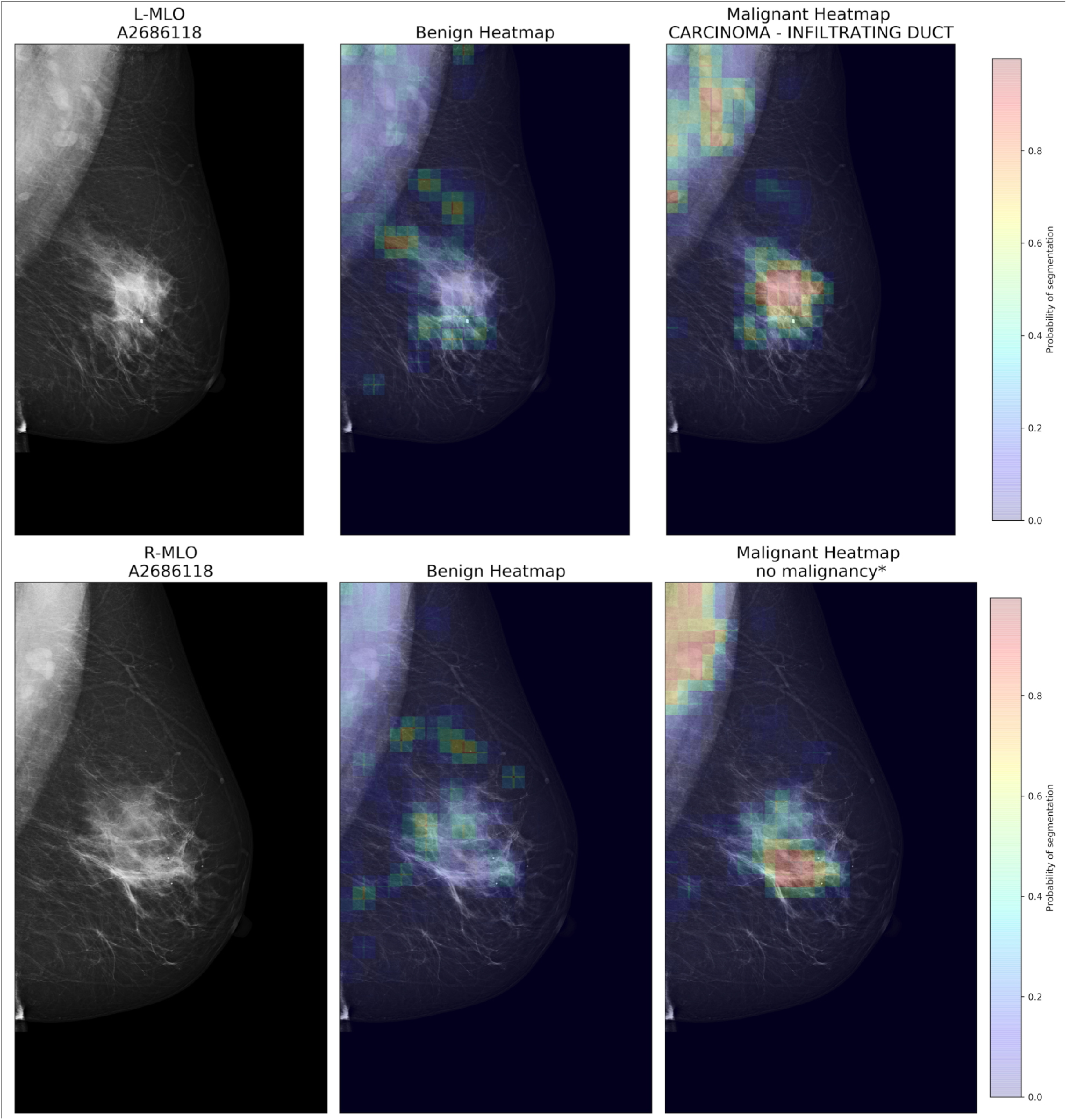
Wu *et al*. heatmaps on BSSA images. Plain MLO mammograms (left) with overlayed benign (middle) and malignant (right) patch-level heatmaps in a single client with biopsy proven invasive ductal carcinoma. These heatmaps are produced by a CNN trained by Wu et. al. (2) (this model was not retrained). Top: left mediolateral oblique view. Bottom: right mediolateral oblique view. Note the high probability of malignant segmentation for densities in the malignancy-free right breast and bilaterally in the axillae. Right-sided images were horizontally flipped for model training, validation and testing. *Malignancy in the contralateral breast.

Wu *et al*. reported on two models, one which used the four-view mammograms only as input (which we refer to as NYU1), and another which included the heatmaps as well as the four-view mammograms (NYU2). Our initial replication analyses focused on applying the original NYU models to local SA data without any retraining on local BSSA data, i.e., static model replication.

### B. Retraining on BSSA Local Data

NYU1 and NYU2 CNNs were retrained on the BSSA training subset with hyperparameter selection using the validation BSSA subsets. The heatmap-generating CNN was not retrained. Code was refactored to utilise pytorch lightning(29), a pytorch wrapper. After selecting the best performing hyperparameters on the validation data (see Supplementary Note 4 - Model Training), we report the number of epochs to the best validation set performance and the AUCs of each model after training “from scratch” with no transfer learning and after retraining with transfer learning (using the NYU weights).

### C. Additional Experiments

We assessed the effect of a lower proportion of malignancy on performance with a second test set (Test Set 2 - see Figure 1 and Table 1). We re-sampled a larger number of age-matched control clients to approximate the prevalence of malignancy in the work by Wu *et al*. (8.4% breast-wise cancer incidence, 16.8% client-wise incidence). Test Set 2 included the same malignant and benign lesion clients as the balanced Test Set 1 and none of the control clients used in any of the training, validation or Test Set 1 subsets. In balancing computational cost, approximating the client-wise prevalence used by the NYU group, and the ‘real-world’ prevalence of malignancy in a screening population, the actual client-wise prevalence in the second test set was 13.06% (see Figure 1).

We further investigated the cumulative effect on AUC of transfer learning and heatmaps, compared to training from scratch with images only, and produced separate results for differentiating malignancy from benign lesions and age-matched controls, stratified by client characteristics and radiological findings.

## Results

For comparison of performance, we refer to the results of the single models described by Wu *et al*. (rather than the ensemble results).(2) The replication analysis of the static NYU1 model achieved an AUC on the local Test Set 1 of 75.8% (95%CI=72.6-78.8%), compared to 83.0% (95%CI=82.4-83.6%) on NYU data, a drop of 7.2% (see Figure 3 and Table 2). The static NYU2 model achieved an AUC of 84.5% (95%CI=81.9-86.8%) on local data, compared to 88.6% (95%CI=88.3%-88.9%) on NYU data(2), a drop of 4.1%. Training of NYU1 and NYU2 on local data, without transfer learning, resulted in AUCs of 65.8% (95%CI=62.2-69.1%) and 85.9 (95%CI=83.5-88.2%), respectively. Retraining of NYU1 and NYU2 on local data, with transfer learning, resulted in modest performance improvements of 6.6% (AUC = 82.4% (95%CI=79.7-84.9%)) and 1.8% (AUC = 86.3% (95%CI=84.0-88.5%)) respectively.

**Table 2.** AUROC for malignancy differentiation. NYU1: image-only models. NYU2: images and benign and malignant heatmaps as model input, pictured in Figure 2 and described by Wu *et al*.(2)

| Malignancy Differentiation on Test Set 1:<br>Area Under the Receiver Operating Characteristic Curve (95% CI) and Epochs |  |  |  |
| --- | --- | --- | --- |
|  | Static model | Trained from scratch | Retrained with transfer learning |
| NYU1 | 75.8 (72.6, 78.8) | 65.8 (62.2 - 69.1)<br>144 epochs | 82.4 (79.7, 84.9)<br>164 epochs |
| NYU2 | 84.5 (81.9, 86.8) | 85.9 (83.5, 88.2)<br>132 epochs | **86.3** (84.0 - 88.5)<br>76 epochs |

**Fig. 3.**
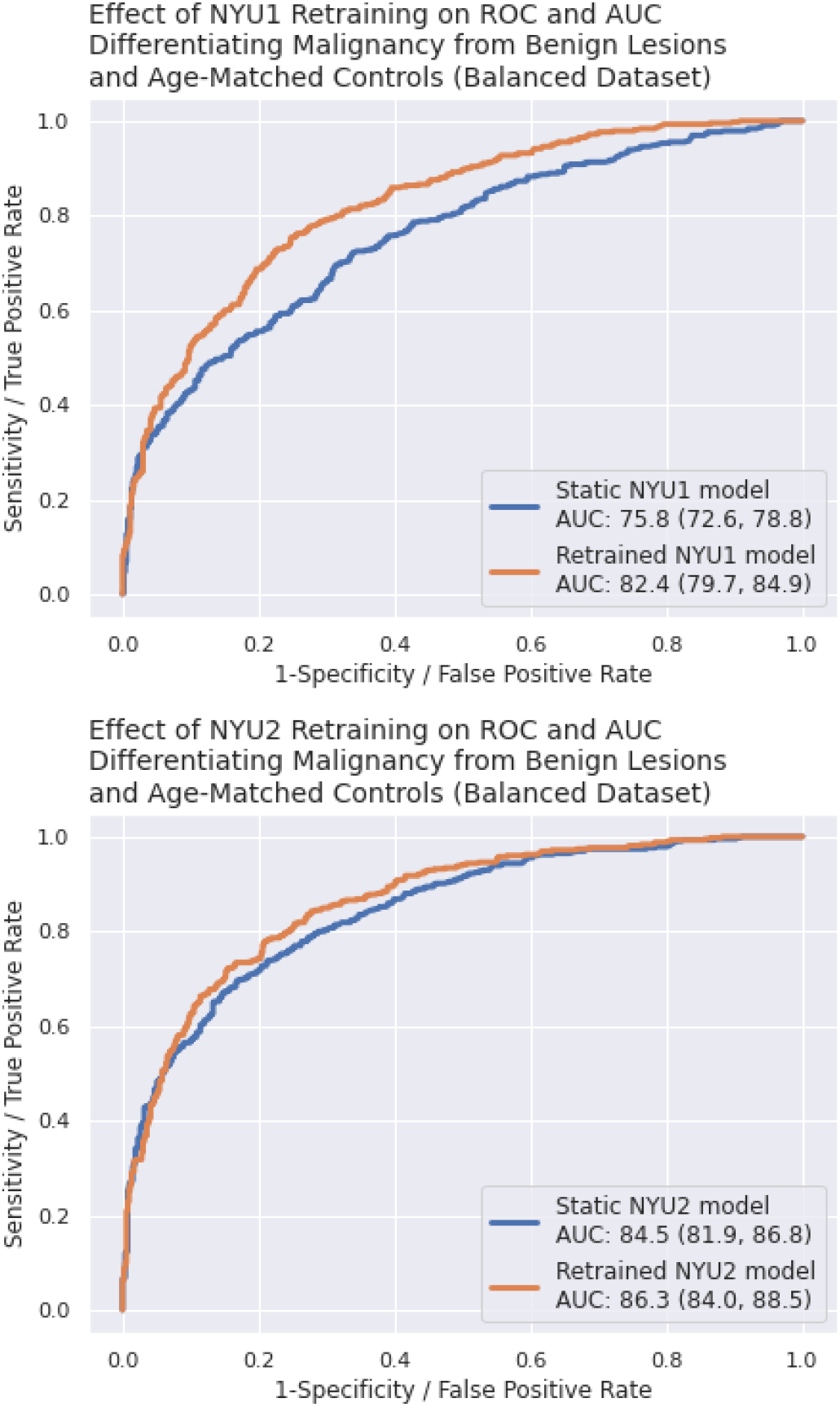
Top: Effect of retraining image-only (NYU1) model. Bottom: Effect of retraining image and heatmaps (NYU2) model.

The NYU2 model retrained with transfer learning achieved an AUC on Test Set 2 of 87.9 (95%CI=86.0-89.6% See Figure S2). For the best model (retrained NYU2), with an operating point chosen for Test Set 1 sensitivity above 90%, the specificity was 59.9%. For results by radiological and client strata, see Table 3 and Supplementary Figure S1.

**Table 3.** NYU2 retrained with transfer learning results by strata - AUROC for differentiating malignancy from benign lesions and age-matched controls in Test Set 1 (except malignant calcification which is calculated against benign calcification). AD: Architectural Distortion

| Category | Subgroup / strata | Clients (n) | AUC (95% CI) |
| --- | --- | --- | --- |
| **Age** |  |  |  |
|  | < 50 | 56 | 90.6 (82.0, 97.2) |
|  | ≥ 50 | 903 | 86.0 (83.6, 88.4) |
| **Subtypes** |  |  |  |
|  | IBC | 340 | 87.5 (85.0, 89.8) |
|  | DCIS | 85 | 90.8 (87.8, 93.5) |
| **Assessment finding** |  |  |  |
|  | Discrete mass | 91 | 87.5 (83.1, 91.5) |
|  | Non-specific density, AD and 'other' | 44 | 81.8 (74.5, 88.1) |
|  | Stellates and multiple masses | 165 | 87.1 (83.8, 90.0) |
|  | Malignant calcification (versus benign calcification) | 142/16 | 77.9 (69.3, 85.7) |
| **Size of invasive malignancy** |  |  |  |
|  | ≤15mm | 311 | 87.9 (85.4, 90.3) |
|  | >15mm | 106 | 88.5 (84.8, 91.8) |
| **Mammography system vendor** |  |  |  |
|  | Philips | 517 | 83.4 (79.8, 86.7) |
|  | Sectra | 442 | 86.2 (82.3, 89.9) |

## Discussion

To our knowledge, this is the first study to apply DL to an Australian mammography dataset and the first to apply an internationally trained DL system to an Australian dataset. We have investigated the replication of open-access deep learning models for screening mammography developed in the USA in data from South Australia. We demonstrate incomplete replication and hence generalisability of the NYU models in local data. Performance and generalisation are likely to improve with the use of larger training datasets.

There was a decrease in diagnostic accuracy when applying the openly available models of Wu *et al*.(2) to the BSSA dataset. By retraining with transfer learning on local data, we were able to achieve a level of performance similar to that originally reported by Wu *et al*. on NYU data. The observed improved performance with retraining was dependent upon the availability of the original model and weights. Without access to weights, retraining with transfer learning is not possible. Replication may depend both on the amount of data used to train the initial model and upon the amount of local data available for retraining, among other factors. Recent external testing shows that some well-trained models do appear to generalise well across populations and mammography system vendors (external testing AUC of 95.6%) (20). Potentially, a sufficiently large number of training examples may improve generalisability to a point where retraining on local data either does not improve performance, or the improvement is negligible. Regardless, our results show that testing of models on local data prior to clinical use is critical to avoid potentially significant decreases in diagnostic accuracy.

There are several possible explanations for the reduced static model performance. There were subtle visual differences when comparing BSSA and NYU images, despite preprocessing (see Figure 4 and Supplementary Note 2). The vendors of imaging equipment used to train the original NYU model were Mammomat Inspiration (22.81%), Mammomat Novation DR (12.65%), Lorad Selenia (40.92%) and Selenia Dimensions (23.62%)(2) contrasting with BSSA equipment vendors Sectra (46.03%) and Philips (53.97%) (see Table 1). Performance variation based on the vendor of the equipment has been described previously(27). Performance across vendors in our dataset was similar, although the different vendors between NYU model training and BSSA may be a source of reduced generalisability. Subtle visual image differences as well as different vendors or sets of vendors between training and clinical deployment datasets may continue to be potential sources of reduced performance in the future.

**Fig. 4.**
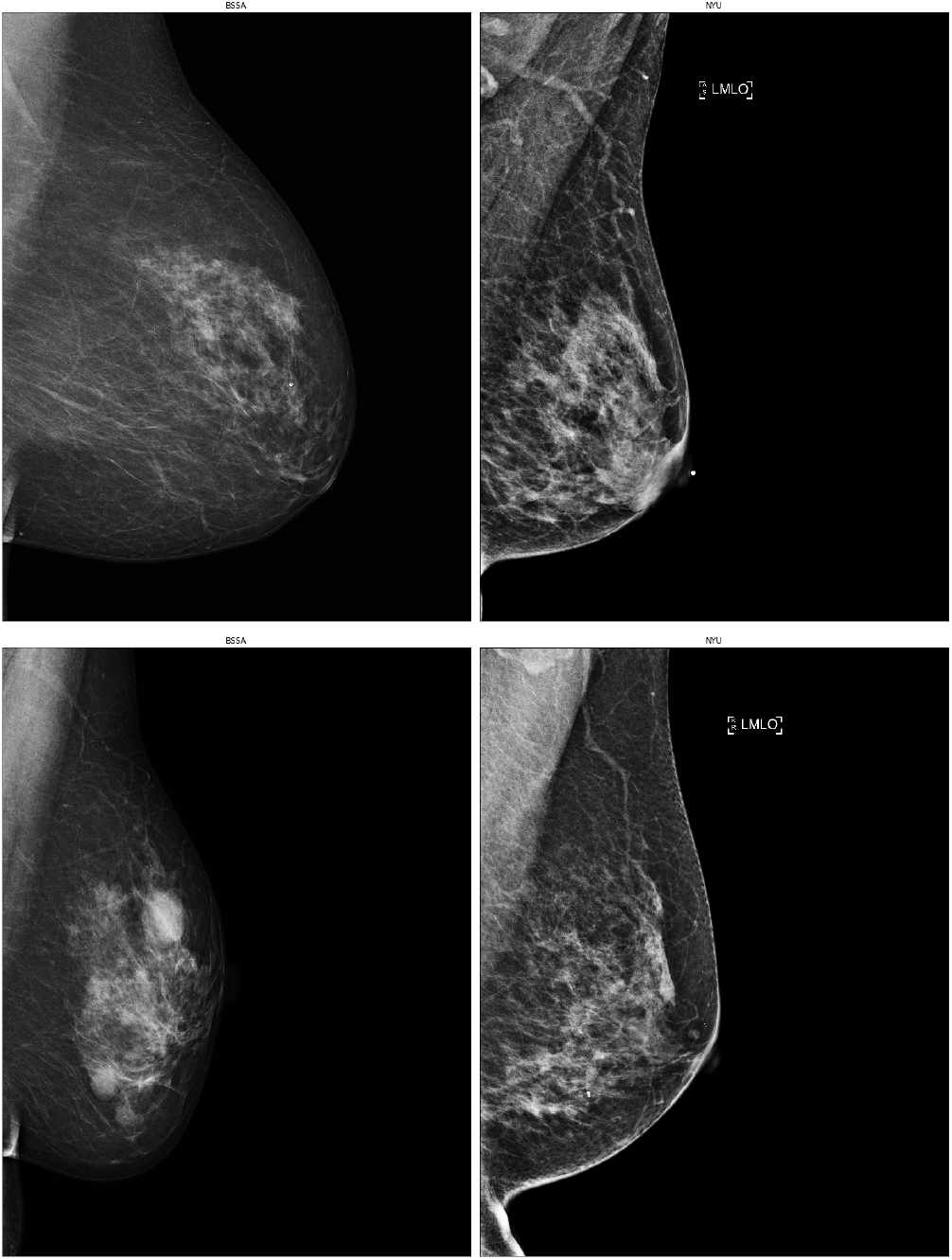
Image differences: BSSA images (left) and NYU images (right) not to scale. Two pairs of MLO views showing subtle differences in image contrast and opacity which may affect model generalisability.

The ground truth may potentially be a source of variation in diagnostic accuracy when attempting to replicate mammography DL systems across populations and datasets. Except for a small number of women who did not undergo surgery, our ground truth for clients with malignancy is the post-surgical specimen where all excised tissue undergoes pathological examination and diagnosis with systematised nomenclature in medicine (SNOMED) classification. The ground-truth in Wu *et al*.(2) was based on text mining of biopsy pathology reports and we also see that a significantly larger portion of NYU biopsies were for benign breasts (76.2% versus 10.2% in the BSSA dataset; see supplementary Table S3). The pathological subtype terminology used by BSSA were different to those presented in Wu et. al. (see supplement Tables S3, S4, S5, S6). In screening mammography datasets, it is possible that women with screening rounds labeled as effectively normal subsequently went on to develop a tumour which was diagnosed and treated in a separate medical system, although this is unlikely to have a large impact on our modelling. Despite differences in ground-truth labelling, both models were ‘ignorant’ of tumour subclass labels and were trained at a higher level of abstraction: malignant or not. While different demographic and lifestyle factors are associated with the prevalence of tumour subtypes (30, 31), the absolute number of subtypes and the magnitude of any differing radiological appearances is unlikely to have any significant effect on performance. In addition, some proportion of invasive cancers will likely be completely mammographically occult, only detectable with other modalities such as ultrasound and MRI (based on current technology). If this proportion differs significantly across populations, this may affect performance of externally tested DL systems. Of note, due to our exclusion criteria, the task of differentiating malignancy in this dataset is less difficult compared to clinical practice. The visual detection of malignancy in clients with only one breast, previous biopsies and/or surgery, for example, can be more challenging.

There is potential for reduced generalisation of diagnostic DL systems due to differences in image size across datasets (see Supplement figure S5) and this may continue to be an issue in the future with different vendors. Some variation in size is unavoidable in mammography. For example, the acquisition process can introduce variation into the size of breast tissue and its contents, relative to the pressure applied to the breast, and there is obviously a natural variation in breast and lesion size. The NYU system, like many CNNs, accepts only a fixed image size and so there are preprocessing choices when developing or validating DL for a range of image and breast sizes. The degree to which apparent breast size should be standardised, if at all, for CNNs is currently unknown (or unpublished). We noticed that despite lower resolution, using images closer to the size of NYU images improved performance. Static NYU model performance on native, full-sized images was reduced compared to images down-scaled by a factor of two, or matched to NYU image size (whilst using the exact same preprocessing method as Wu *et al*. (excluding initial inversion of images)). See Supplementary Figure S4.

Work by Wu *et al*. (2) included pretraining on over 100,000 clients, using Breast Imaging and Reporting Data System (BIRADS) categories as the target output. This pretraining ran for just under 2 weeks continuously (326 hours), on four Nvidia V100 GPUs. The Wu *et al*. exam-level CNN was trained for 12 hours on an Nvidia V100GPU(2). Benign and malignant heatmaps (see Figure 2), which in the current study improved diagnostic accuracy, were generated as part of the Wu *et al*. freely available system,(1) including a model trained on 4,013 segmentations hand-drawn by NYU radiologists. The sharing of this time-consuming work was of considerable benefit in our experiments when building a “local” model, with retraining taking significantly less time than training from scratch on local data (see Table 2). We also note that retraining using the NYU weights was far quicker than the development of the original Wu *et al*. system (2), despite using much less powerful hardware. In this way, the sharing of models, weights and code expedites research in the field. Recent works suggest that some combined modelling of fine-grained, lesion-level and larger-scale information is superior to modelling whole mammogram images alone. Strategies include using combinations of client-level, image-level and patch-level models (17), pretraining on lesion patches(18) and including patch-level heatmaps as input.(2) The importance of both this fine-grained information and the use of pretrained weights is highlighted in the drop in AUC observed with the NYU1 model trained from scratch, with randomly initialised weights.

Unfortunately, we were not able to assess performance with respect to breast density, as it is not routinely collected in Australia.(32)

### Future Work

We plan to explore how DL systems might integrate with human reader assessments, and the effect this would have on overall screening recall rates, positive predictive value, sensitivity and specificity.

There are several recent studies which include or relate to accommodating for subtle image differences. Recent approaches include an encoder-decoder module at the top of a CNN(18), batch-instance normalisation (18) and neural style transfer(33). This may be significant in the event that a screening service changes equipment vendors, requiring transition of an existing model to slightly different images. In addition, a method to effectively combine large datasets, potentially internationally, is with distributed or ‘federated’ deep learning (34). This may enable the training of large models with images from multiple different sites, whilst client data remains at its original site (35).

## Conclusion

We were not able to fully reproduce the reported performance of Wu *et al*. (2) in an Australian screening mammography dataset. The availability of weights of the NYU models allowed local retraining with transfer learning; model performance in the local SA dataset then approached the levels originally reported. The additional input of pixel-level heatmaps improved performance, even without local retraining of the heatmap generating model, supporting the findings reported in Wu *et al*.(2) Shared models and code enabled our experiments, including to provide independent external testing of the system, to test the effects of including heatmaps as model inputs, and to investigate the effects of local retraining. Our findings suggest that (1) replication of initial model performance in different clinical environments may be challenging and (2) access to model weights may be necessary to enable datasets to be adapted to local clinical environments. Local testing of DL systems is likely to be necessary for pre-deployment model assessment, regulatory approval, and safe clinical translation. Reduced performance of DL systems in new environments may create challenges to the widespread use of DL in radiology.

## Data Availability

The BSSA dataset is private and not able to be released. The data that support the results presented will be made available at www.github.com/jamesjjcondon

## Author Contributions

AH managed and quality assured the extraction of non-dicom client data. KAH wrote SQL scripts to extract client data from the BSSA in-house database and upload to secure storage. JJC wrote and executed all training, inference and testing code for this project (excluding that provided by Wu *et al*.(1) JJC and LOR examined image samples for effects of preprocessing including LUT transforms and downscaling. JJC, LOR, LJP and GC designed the study and analysed the results. JJC, LOR, KAR, AH, MR, GC and LJP contributed to and reviewed the manuscript.

## Code Availability

All code used for this study will be made available at github.com/jamesjjcondon/breast_cancer_classifier_RT. This repository is not currently operational for replication and relies on the private BSSA data. It is made available for transparency of model retraining and evaluation.

## Acknowledgements

Thanks to the research of Wu, Park, Geras *et al*. and for making their model, weights and code openly available, without which the present work would not be possible. Thanks to Ada Childs for help with extracting the client related data, Jason Chatterton and Daniel Pringle for their work extracting the dicom images. Thanks to Stephan Lau for his advice regarding runtime optimisation. Thanks to William Falcoln, founder of pytorch lightning for this package and technical support. Thanks to countless open-source contributors for the Ubuntu OS and numerous packages on which the project is dependent (pytorch, pydicom, numpy, pandas, h5py, scikit-learn and others).

## Supplementary Note 1: BreastScreen South Australia Dataset

All full-field digital screening and assessment mammograms, were transferred to secure research storage (n images=2 004 985). Only screening images with a view of craniocaudal (CC) or mediolateral oblique (MLO) were used (see main article, Figure 1). The total number of clients with biopsy or surgical pathology-proven lesions in this study was 3160. There were 2837 clients with malignant and 323 with benign lesions. A total of 9185 clients were used in this study although counts vary with respect to inclusion of the balanced Test Set 1 and approximated NYU Test Set 2 (see Table 1. Details of image manufacturers, age, hormone replacement therapy are described in Table 1. Histological diagnoses, tumour stage and their differences across NYU and BSSA are described in Supplementary Tables S1, S2 and S3.

## Supplementary Note 2: Dicoms, Transfer and Image Preparation

Clients’ identities were removed from all dicoms (digital communication in medicine file format) prior to transfer from BSSA to our secure research storage. To enable the transfer of approx 100TBs of data, dicoms underwent lossless compression at BSSA. Dicoms were extracted from the BSSA client Archiving and Communication System. SQL scripts were written to extract client data. Dicoms and client data were then downloaded to physical external hard drives. All subsequent model development and analysis used python 3.6.(1) Compressed dicoms were later decompressed with pydicom version 1.3.0.(2)

Mammograms from BSSA were all monochrome 1 photometric interpretation (where pixel array values are low for high density / low x-ray beam intensity and high for low density / high x-ray beam intensity, see figure 1) and used a sigmoid voxel of interest look-up table (LUT). Monochrome 1 images were excluded from Wu’s model development (3), a potential source of reduced generalisation. Dicom viewers may internally process dicom metadata and pixel arrays into a presentation state for radiologists. For monochrome 1 photometric interpretation, this is achieved with a standard algorithm:

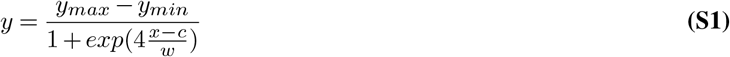

where ‘ymax’ and ‘ymin’ are the maximum and minimum output values, ‘x’ is raw pixel value, ‘c’ is the ‘normal’ window centre and ‘w’ is the ‘normal’ window width (4, 5). This maps raw pixel values to presentation state output with respect to a) the pixel intensity relationship b) voxels of Interest (VOI) look-up table (LUT) function and c) four possible tuples of contrast window centre and width (eg ‘User’ (defined), ‘Bright’, ‘Normal’ or ‘Dark’). BSSA dicoms consisted of a sigmoid VOI LUT function, log pixel intensity relationship and an inverse presentation LUT shape. We wrote a function to apply this algorithm so that images were converted to a presentation state. This facilitated both ‘sanity checks’ of images and we felt was more likely to improve generalisability (versus using inverted images). See Figure S4 for visualisation of pixel arrays before and after conversion.

Unsigned 16-bit integer dicom pixel arrays were converted to 32-bit, necessarily, to match NYU weights. Our images consisted of a 5355 × 4915 pixel array for both CC and MLO views. After testing the effect of downscaling on the unchanged NYU model, images were downscaled with bicubic interpolation to match the smallest NYU sample image height (3328) and width selected to preserve aspect ratio, at the beginning of preprocessing with minor adjustments to original code. Using code from (6), an ‘ideal centre’ of the image is calculated during preprocessing, upon which crop augmentations are based for training, validation and testing augmentation.

## Supplementary Note 3: Histogram Matching

We attempted to remove domain effects and improve unchanged NYU model performance by matching BSSA images to histograms of sample data openly provided by Wu et al (3, 6). Some authors have used this to account for variation in pixel values.(8) Scikit-image (7) source code was patched to calculate and match histograms using non-zero values only. NYU sample data consists of 4 exams (16 images). During preprocessing BSSA images (the source image in Figure S6) were matched with one of the 8 corresponding sampled images of the same view (CC or MLO) however this resulted in significant degradation of performance.

**Table S1.**
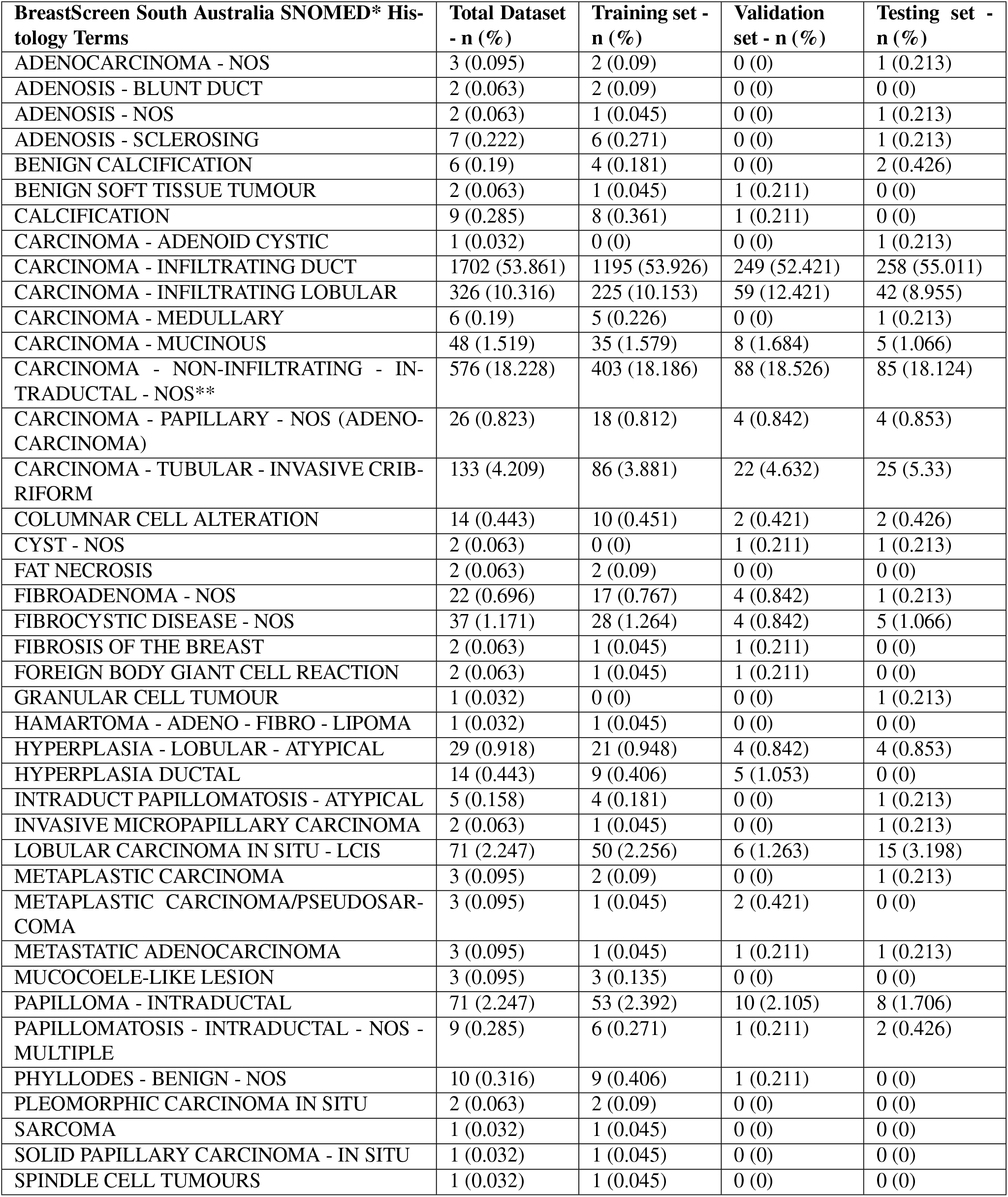
All biopsy-proven benign and malignant lesions BSSA Jan 1 2010 through Dec 31 2016 (n=3160) *Systematised Nomenclature of Medicine. **Equivalent to DCIS

| **BreastScreen South Australia SNOMED\* Histology Terms** | **Total Dataset - n (%)** | **Training set - n (%)** | **Validation set - n (%)** | **Testing set - n (%)** |
| --- | --- | --- | --- | --- |
| ADENOCARCINOMA - NOS | 3 (0.095) | 2 (0.09) | 0 (0) | 1 (0.213) |
| ADENOSIS - BLUNT DUCT | 2 (0.063) | 2 (0.09) | 0 (0) | 0 (0) |
| ADENOSIS - NOS | 2 (0.063) | 1 (0.045) | 0 (0) | 1 (0.213) |
| ADENOSIS - SCLEROSING | 7 (0.222) | 6 (0.271) | 0 (0) | 1 (0.213) |
| BENIGN CALCIFICATION | 6 (0.19) | 4 (0.181) | 0 (0) | 2 (0.426) |
| BENIGN SOFT TISSUE TUMOUR | 2 (0.063) | 1 (0.045) | 1 (0.211) | 0 (0) |
| CALCIFICATION | 9 (0.285) | 8 (0.361) | 1 (0.211) | 0 (0) |
| CARCINOMA - ADENOID CYSTIC | 1 (0.032) | 0 (0) | 0 (0) | 1 (0.213) |
| CARCINOMA - INFILTRATING DUCT | 1702 (53.861) | 1195 (53.926) | 249 (52.421) | 258 (55.011) |
| CARCINOMA - INFILTRATING LOBULAR | 326 (10.316) | 225 (10.153) | 59 (12.421) | 42 (8.955) |
| CARCINOMA - MEDULLARY | 6 (0.19) | 5 (0.226) | 0 (0) | 1 (0.213) |
| CARCINOMA - MUCINOUS | 48 (1.519) | 35 (1.579) | 8 (1.684) | 5 (1.066) |
| CARCINOMA - NON-INFILTRATING - INTRADUCTAL - NOS** | 576 (18.228) | 403 (18.186) | 88 (18.526) | 85 (18.124) |
| CARCINOMA - PAPILLARY - NOS (ADENOCARCINOMA) | 26 (0.823) | 18 (0.812) | 4 (0.842) | 4 (0.853) |
| CARCINOMA - TUBULAR - INVASIVE CRIBRIFORM | 133 (4.209) | 86 (3.881) | 22 (4.632) | 25 (5.33) |
| COLUMNAR CELL ALTERATION | 14 (0.443) | 10 (0.451) | 2 (0.421) | 2 (0.426) |
| CYST - NOS | 2 (0.063) | 0 (0) | 1 (0.211) | 1 (0.213) |
| FAT NECROSIS | 2 (0.063) | 2 (0.09) | 0 (0) | 0 (0) |
| FIBROADENOMA - NOS | 22 (0.696) | 17 (0.767) | 4 (0.842) | 1 (0.213) |
| FIBROCYSTIC DISEASE - NOS | 37 (1.171) | 28 (1.264) | 4 (0.842) | 5 (1.066) |
| FIBROSIS OF THE BREAST | 2 (0.063) | 1 (0.045) | 1 (0.211) | 0 (0) |
| FOREIGN BODY GIANT CELL REACTION | 2 (0.063) | 1 (0.045) | 1 (0.211) | 0 (0) |
| GRANULAR CELL TUMOUR | 1 (0.032) | 0 (0) | 0 (0) | 1 (0.213) |
| HAMARTOMA - ADENO - FIBRO - LIPOMA | 1 (0.032) | 1 (0.045) | 0 (0) | 0 (0) |
| HYPERPLASIA - LOBULAR - ATYPICAL | 29 (0.918) | 21 (0.948) | 4 (0.842) | 4 (0.853) |
| HYPERPLASIA DUCTAL | 14 (0.443) | 9 (0.406) | 5 (1.053) | 0 (0) |
| INTRADUCT PAPILLOMATOSIS - ATYPICAL | 5 (0.158) | 4 (0.181) | 0 (0) | 1 (0.213) |
| INVASIVE MICROPAPILLARY CARCINOMA | 2 (0.063) | 1 (0.045) | 0 (0) | 1 (0.213) |
| LOBULAR CARCINOMA IN SITU - LCIS | 71 (2.247) | 50 (2.256) | 6 (1.263) | 15 (3.198) |
| METAPLASTIC CARCINOMA | 3 (0.095) | 2 (0.09) | 0 (0) | 1 (0.213) |
| METAPLASTIC CARCINOMA/PSEUDOSARCOMA | 3 (0.095) | 1 (0.045) | 2 (0.421) | 0 (0) |
| METASTATIC ADENOCARCINOMA | 3 (0.095) | 1 (0.045) | 1 (0.211) | 1 (0.213) |
| MUCOCOELE-LIKE LESION | 3 (0.095) | 3 (0.135) | 0 (0) | 0 (0) |
| PAPILLOMA - INTRADUCTAL | 71 (2.247) | 53 (2.392) | 10 (2.105) | 8 (1.706) |
| PAPILLOMATOSIS - INTRADUCTAL - NOS - MULTIPLE | 9 (0.285) | 6 (0.271) | 1 (0.211) | 2 (0.426) |
| PHYLLODES - BENIGN - NOS | 10 (0.316) | 9 (0.406) | 1 (0.211) | 0 (0) |
| PLEOMORPHIC CARCINOMA IN SITU | 2 (0.063) | 2 (0.09) | 0 (0) | 0 (0) |
| SARCOMA | 1 (0.032) | 1 (0.045) | 0 (0) | 0 (0) |
| SOLID PAPILLARY CARCINOMA - IN SITU | 1 (0.032) | 1 (0.045) | 0 (0) | 0 (0) |
| SPINDLE CELL TUMOURS | 1 (0.032) | 1 (0.045) | 0 (0) | 0 (0) |

**Table S2.**
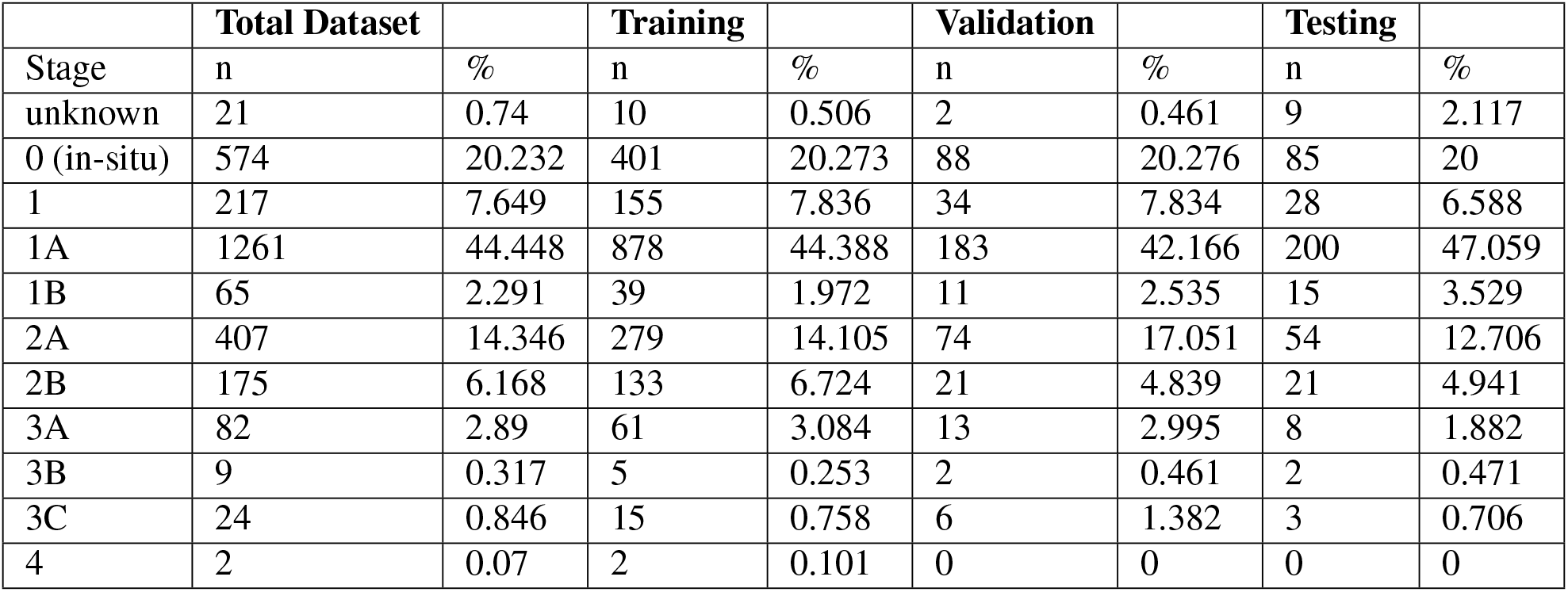
Tumour Stage for dataset and subsets

**Table S3.** Comparing breast biopsies and ground truth labels

|  | BSSA | NYU |
| --- | --- | --- |
| Total Malignant Breasts | 2837 | 985 |
| Total Benign Breasts | 323 | 3158 |
| Malignant breasts: All biopsied breasts (%) | 0.898 | 0.238 |
| Benign breasts: All biopsied breasts (%) | 0.102 | 0.762 |
| Ground truth provenance | Post-surgical or biopsy SNOMED classification | Text extraction, classification and grouping from biopsy pathology reports |

**Table S4.** Malignant pathology terms BSSA versus NYU

| BSSA | NYU |
| --- | --- |
| "ADENOCARCINOMA - NOS" | "adenocarcinoma" |
| "CARCINOMA - ADENOID CYSTIC" | "ductal carcinoma" |
| "CARCINOMA - INFILTRATING DUCT" | "ductal carcinoma in situ" |
| "CARCINOMA - INFILTRATING LOBULAR" | "invasive carcinoma" |
| "CARCINOMA - MEDULLARY" | "invasive ductal carcinoma" |
| "CARCINOMA - MUCINOUS" | "invasive lobular carcinoma" |
| "CARCINOMA - NON-INFILTRATING - INTRADUCTAL - NOS" | "invasive mammary carcinoma" |
| "CARCINOMA - PAPILLARY - NOS (ADENOCARCINOMA)" | "metastases" |
| "CARCINOMA - TUBULAR - INVASIVE CRIBIFORM" | "metastatic" |
| "INVASIVE MICROPAPILLARY CARCINOMA" | "metastatic carcinoma" |
| "METAPLASTIC CARCINOMA" |  |
| "METAPLASTIC CARCINOMA/PSEUDOSARCOMA" |  |
| "METASTATIC ADENOCARCINOMA" |  |
| "PLEOMORPHIC CARCINOMA IN SITU" |  |
| "SARCOMA" |  |
| "SOLID PAPILLARY CARCINOMA - IN SITU" |  |
| "SPINDLE CELL TUMOURS" |  |

**Table S5.** Benign pathology terms BSSA versus NYU

| BSSA | NYU |
| --- | --- |
| "ADENOSIS - BLUNT DUCT"<br>"ADENOSIS - FLORID"<br>"ADENOSIS - NOS"<br>"ADENOSIS - SCLEROSING"<br>"BENIGN CALCIFICATION"<br>"BENIGN SOFT TISSUE TUMOUR"<br>"CALCIFICATION"<br>"COLUMNAR CELL ALTERATION"<br>"CYST - NOS"<br>"FAT NECROSIS"<br>"FIBROADENOMA - NOS"<br>"FIBROCYSTIC DISEASE - NOS"<br>"FIBROSIS OF THE BREAST"<br>"FOREIGN BODY GIANT CELL REACTION"<br>"GRANULAR CELL TUMOUR"<br>"HAMARTOMA - ADENO - FIBRO - LIPOMA"<br>"HYPERPLASIA - LOBULAR - ATYPICAL"<br>"HYPERPLASIA DUCTAL"<br>"INTRADUCT PAPILLOMATOSIS - ATYPICAL"<br>"LOBULAR CARCINOMA IN SITU - LCIS"<br>"MUCOCOELE-LIKE LESION"<br>"PAPILLOMA - INTRADUCTAL"<br>"PAPILLOMATOSIS - INTRADUCTAL - NOS - MULTIPLE"<br>"PHYLLODES - BENIGN - NOS" | "adipose tissue"<br>"benign breast tissue"<br>"cyst content"<br>"fibroadenoma"<br>"fibrocystic change"<br>"fibrocystic changes"<br>"fibrosis"<br>"hyperplasia" |

**Table S6.** Exclusion pathology terms BSSA versus NYU

| BSSA | NYU |
| --- | --- |
| "ECTASIA - MAMMARY DUCT - PLASMA CELL MASTITIS"<br>"GRANULOMATOUS INFLAMMATION"<br>"INFLAMMATION - CHRONIC - NOS"<br>"MASTITIS"<br>"METAPLASIA - APOCRINE"<br>"NON-SPECIFIC REACTIVE PROCESS"<br>"NORMAL TISSUE - NOS"<br>"PHYLLODES - BORDERLINE - NOS"<br>"RADIAL SCAR"<br>"UNKNOWN" | "negative for malignancy"<br>"benign skin"<br>"breast capsule"<br>"breast implant"<br>"dermal scar"<br>"explant"<br>"fibrous capsule"<br>"no benign or malignant epithelial cells seen"<br>"no mammary epithelial cells"<br>"no mammary epithelium identified"<br>"non-diagnostic" |

## Supplementary Note 4: Model Training

During training, four views for each client were fed to the model (left and right CC and MLO), with random selection from multiple breast views, if they existed, as in (6). As described by Wu *et al*., the client-level CNN (Resnet-22) model accepts a dictionary of four images with separate fully connected layers for CC and MLO. Left and right features for each view are concatenated before entering the respective fully connected layer.

Each image was normalised to a mean of zero and standard deviation of one during augmentation as in (3, 6). Augmentation during training was aligned with that used in Wu et al in order to maximise transfer learning. Code from (6) adds gaussian noise with a standard deviation of 0.01 to all images during training and is removed for validation and testing.

We used distributed data parallelism (where each of 2 GPUs hosts a replica of the model and trains on the entire dataset with synchronisation of weight updates) as empirically it is more computationally efficient then data parallelism. There are significant overheads of CPU-GPU communication when using large images. Learning rate was reduced by a factor of 0.1 when it plateaued with a minimum of 1e-12, patience of two epochs and default parameters for base momentum and maximum momentum (0.8 and 0.9 respectively).

Negative log likelihood loss was used as the objective function, accepting 8 model output log probabilities (one for each benign and malignant class for each view per breast). This is equivalent to binary cross entropy loss, with a separate loss calculated for each view for each of benign and malignant findings as in (6). Weight updates were calculated with the Adam optimiser (9) with L2 regularisation weight decay of 4e-05. Mixed precision was achieved with the python-only build of NVIDIA’s apex pytorch extension (10), enabling a batch size of 6 (where with 32-bit precision, only 4 would fit on our 12GB cards). Gradients were accumulated for 6 batches, achieving an effective batch size of 36. Learning curves (training and validation loss and accuracy) were monitored with tensorboard and training manually terminated.

## Supplementary Note 5: Inference

Window cropping during inference may result in slightly different outputs for each iteration, with the average of 10 iterations per client used, as in (6). Softmax scores for benign and malignant breast are averaged for each view to calculate breast-wise scores. The maximum score for benign and malignant lesions across both breasts were used for client-wise scores.

We experimented with additional augmentation at test time consisting of horizontal and vertical flips and rotation (of a random degree 0-360) each with a probability of 0.5, which some have found improves performance (11) however performance decreased slightly (presumably because this was never part of training augmentations).

The final models for testing were selected based on the nadir of validation loss. Apart from pure code debugging, inference on the test set was run once. The test set was not used for any form of validation, model training or hyperparameter optimisation.

---

**Fig. S1.**
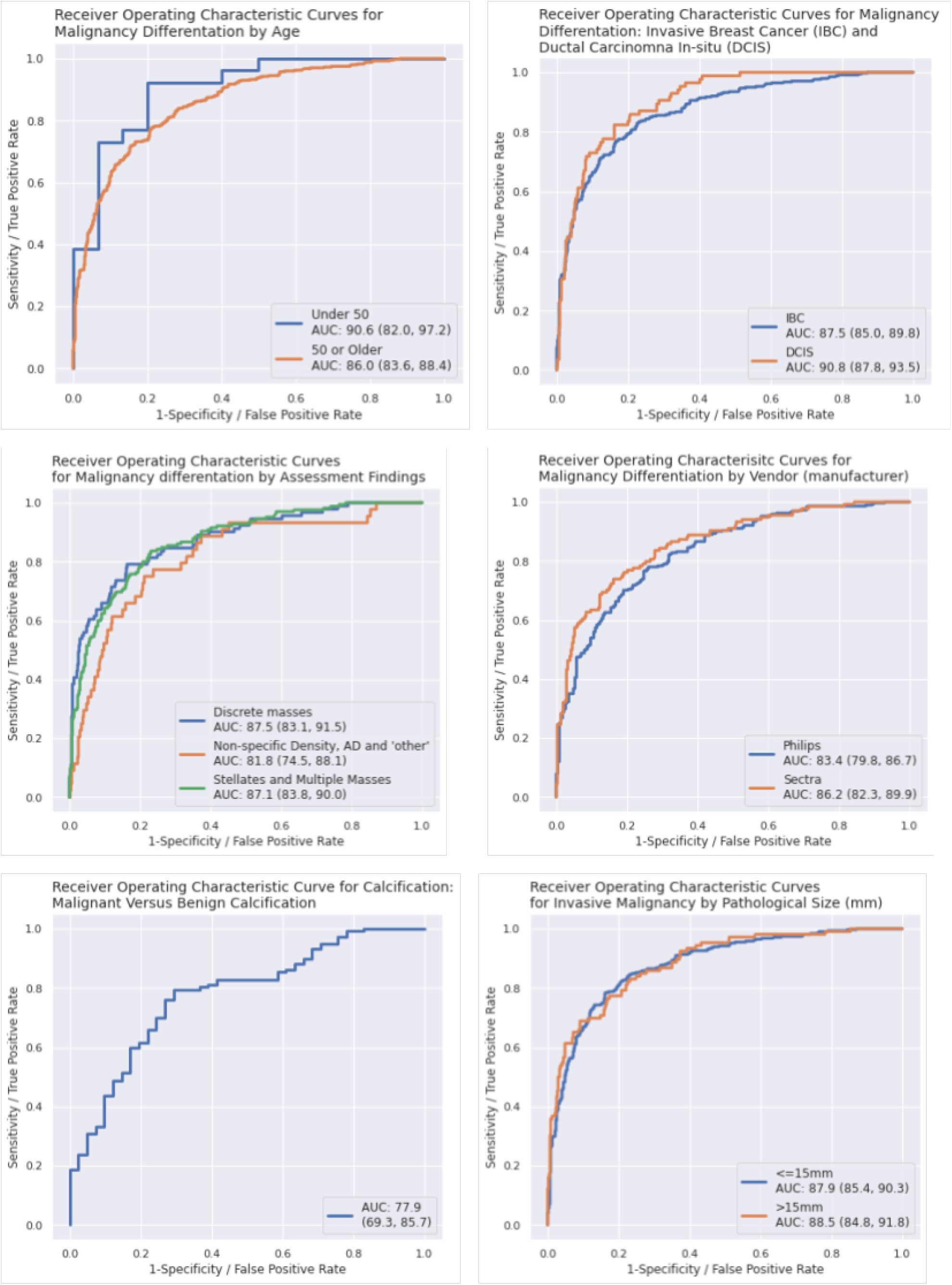
Secondary results by client and radiological strata for NYU2 retrained with transfer learning.

**Fig. S2.**
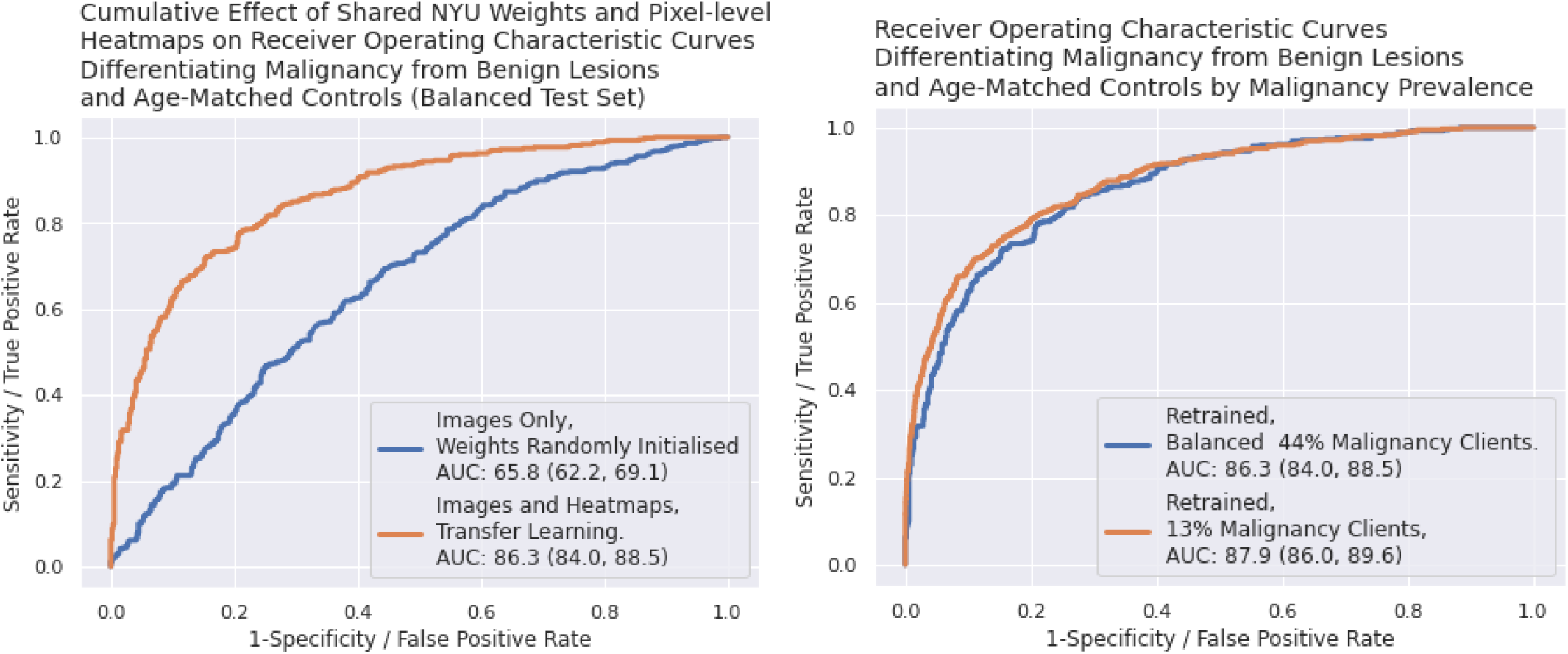
Left: cumulative effect of NYU heatmaps and weights. Right: AUROC by malignancy incidence

**Fig. S3.**
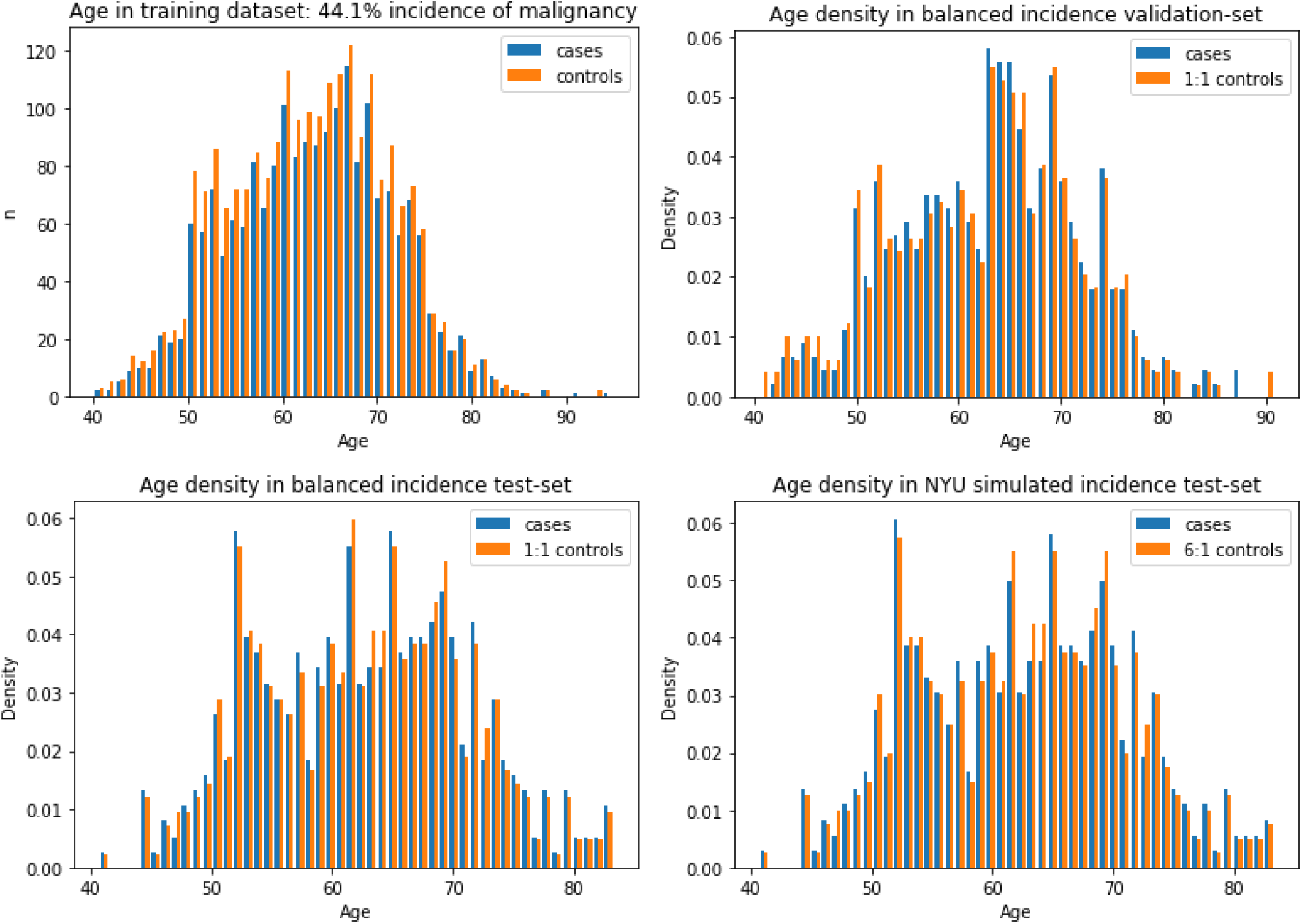
Age histograms for training, validation, balanced test set 1, and imbalanced test set 2

**Fig. S4.**
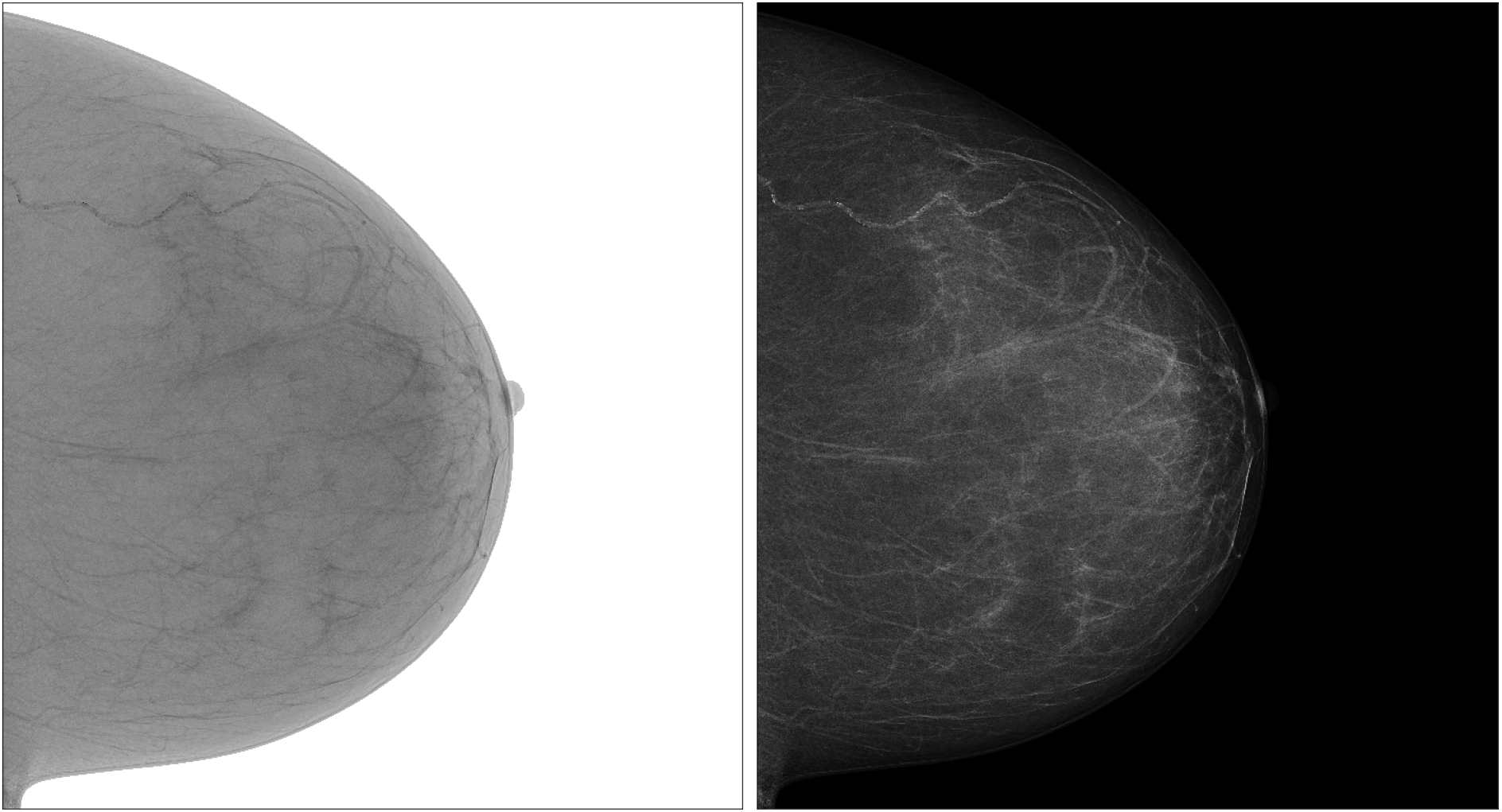
Transformation from inverted monochrome 1 to presentation state.

**Fig. S5.**
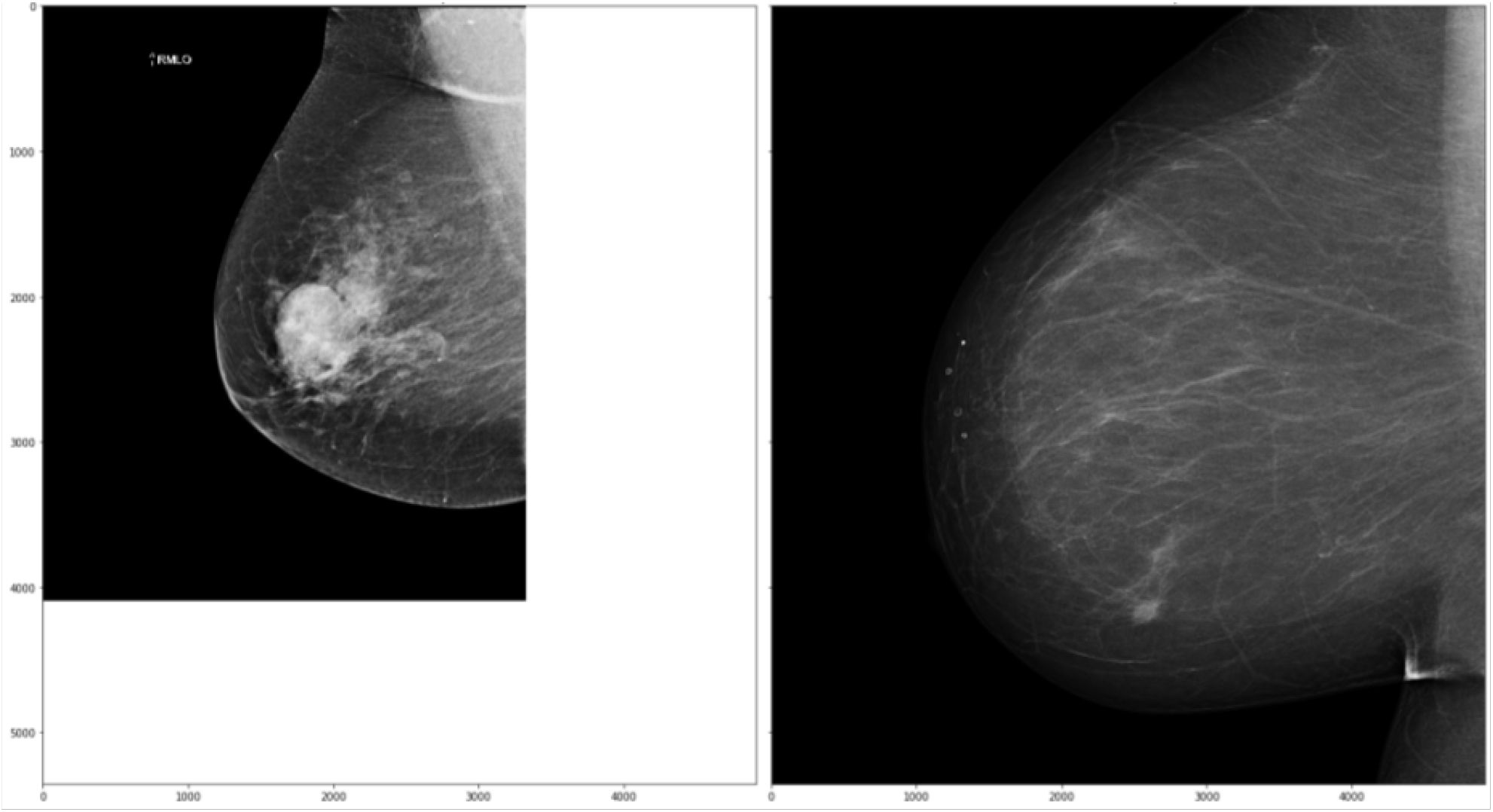
Different image sizes to scale. Left: NYU image - 4096 × 3328 pixels. Right: BSSA image - 5355 × 4915 pixels

**Fig. S6.**
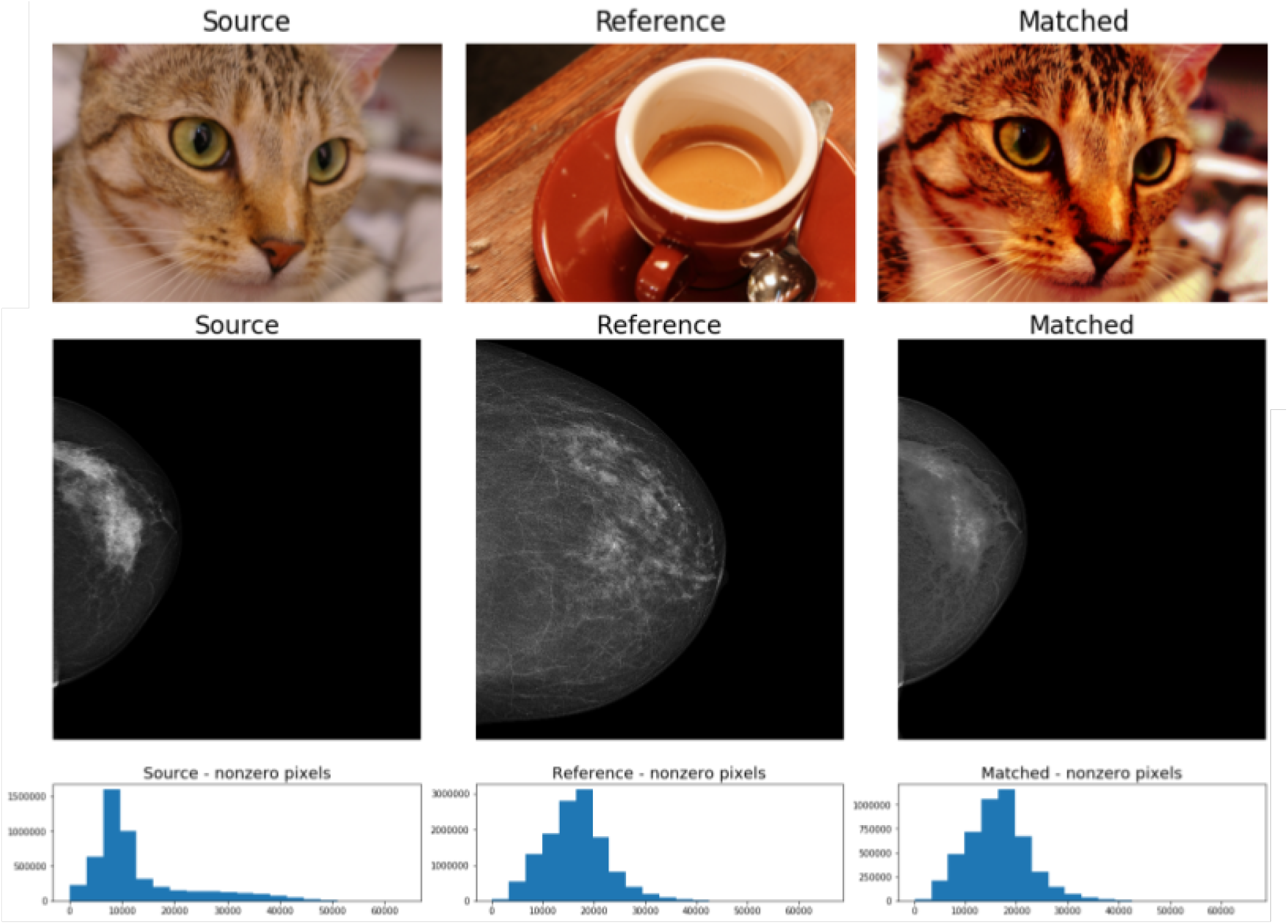
Histogram matching (7). Columns left to right: source image including BSSA CC example; reference image including NYU example sample image (6); source imaged matched to non-zero histogram of reference image.

## References

1. nyukat. nyukat/breast_cancer_classifier. https://github.com/nyukat/breast_cancer_classifier. Accessed: 2020-2-9.

2. Wu N, Phang J, Park J et al. Deep neural networks improve radiologists’ performance in breast cancer screening. - PubMed - NCBI. https://www.ncbi.nlm.nih.gov/ pubmed/31603772. Accessed: 2020-3-6.

3. BreastScreen australia monitoring report 2014–2015, table of contents - australian institute of health and welfare. https://www.aihw.gov.au/reports/ cancer-screening/breastscreen-australia-monitoring-2014-15/contents/table-of-contents,. Accessed: 2019-6-30.

4. Sepideh Saadatmand, Reini Bretveld, Sabine Siesling, and Madeleine M A Tilanus-Linthorst. Influence of tumour stage at breast cancer detection on survival in modern times: population based study in 173 797 patients, 2015.

5. S Rossi, C Cinini, C Di Pietro, C P Lombardi, A Crucitti, R Bellantone, and F Crucitti. Diagnostic delay in breast cancer: correlation with disease stage and prognosis. Tumori, 76 (6):559–562, December 1990.

6. Theofilou Paraskevi. Quality of life outcomes in patients with breast cancer, 2012.

7. D Roder, N Houssami, G Farshid, G Gill, C Luke, P Downey, K Beckmann, P Iosifidis, L Grieve, and L Williamson. Population screening and intensity of screening are associated with reduced breast cancer mortality: evidence of efficacy of mammography screening in australia. Breast Cancer Res. Treat., 108(3), April 2008.

8. P Strax, L Venet, and S Shapiro. Value of mammography in reduction of mortality from breast cancer in mass screening. Am. J. Roentgenol. Radium Ther. Nucl. Med., 117(3): 686–689, March 1973.

9. László Tabár, Bedrich Vitak, Tony Hsiu-Hsi Chen, Amy Ming-Fang Yen, Anders Cohen, Tibor Tot, Sherry Yueh-Hsia Chiu, Sam Li-Sheng Chen, Jean Ching-Yuan Fann, Johan Rosell, Helena Fohlin, Robert A Smith, and Stephen W Duffy. Swedish Two-County trial: Impact of mammographic screening on breast cancer mortality during 3 decades, 2011.

10. Lennarth Nyström, Ingvar Andersson, Nils Bjurstam, Jan Frisell, B. Nordenskjöld, and Lars Erik Rutqvist. Long-term effects of mammography screening: updated overview of the swedish randomised trials, 2002.

11. Nils Bjurstam, Lena Björneld, Jane Warwick, Evis Sala, Stephen W Duffy, Lennarth Nyström, Neil Walker, Erling Cahlin, Olof Eriksson, Lars-Olof Hafström, Halvard Lingaas, Jan Mattsson, Stellan Persson, Carl-Magnus Rudenstam, Håkan Salander, Johan Säve-Söderbergh, and Torkel Wahlin. The gothenburg breast screening trial, 2003.

12. Sue M Moss, Christopher Wale, Robert Smith, Andrew Evans, Howard Cuckle, and Stephen W Duffy. Effect of mammographic screening from age 40 years on breast cancer mortality in the UK age trial at 17 years’ follow-up: a randomised controlled trial, 2015.

13. BreastScreen australia data dictionary: version 1.2. https://www.aihw.gov.au/reports/cancer-screening/breastscreen-australia-data-dictionary-version-1-2/contents/ table-of-contents,. Accessed: 2021-4-26.

14. Constance D Lehman, Robert D Wellman, Diana SM Buist, Karla Kerlikowske, Anna NA Tosteson, and Diana L Miglioretti. Diagnostic accuracy of digital screening mammography with and without computer-aided detection. JAMA internal medicine, 175(11):1828–1837, 2015.

15. Yann LeCun, Yoshua Bengio, and Geoffrey Hinton. Deep learning, 2015.

16. S. J. Pan and Q. Yang. A survey on transfer learning. IEEE Transactions on Knowledge and Data Engineering, 22(10):1345–1359, 2010. doi: 10.1109/TKDE.2009.191.

17. Scott Mayer McKinney, Marcin Sieniek, Varun Godbole, Jonathan Godwin, Natasha Antropova, Hutan Ashrafian, Trevor Back, Mary Chesus, Greg C Corrado, Ara Darzi, Mozziyar Etemadi, Florencia Garcia-Vicente, Fiona J Gilbert, Mark Halling-Brown, Demis Hassabis, Sunny Jansen, Alan Karthikesalingam, Christopher J Kelly, Dominic King, Joseph R Ledsam, David Melnick, Hormuz Mostofi, Lily Peng, Joshua Jay Reicher, Bernardino Romera-Paredes, Richard Sidebottom, Mustafa Suleyman, Daniel Tse, Kenneth C Young, Jeffrey De Fauw, and Shravya Shetty. International evaluation of an AI system for breast cancer screening. Nature, 577(7788):89–94, January 2020.

18. Hyo-Eun Kim, Hak Hee Kim, Boo-Kyung Han, Ki Hwan Kim, Kyunghwa Han, Hyeonseob Nam, Eun Hye Lee, and Eun-Kyung Kim. Changes in cancer detection and false-positive recall in mammography using artificial intelligence: a retrospective, multireader study, 2020.

19. Alejandro Rodríguez-Ruiz, Elizabeth Krupinski, Jan-Jurre Mordang, Kathy Schilling, Sylvia H Heywang-Köbrunner, Ioannis Sechopoulos, and Ritse M Mann. Detection of breast cancer with mammography: Effect of an artificial intelligence support system, 2019.

20. Mattie Salim, Erik Wåhlin, Karin Dembrower, Edward Azavedo, Theodoros Foukakis, Yue Liu, Kevin Smith, Martin Eklund, and Fredrik Strand. External evaluation of 3 commercial artificial intelligence algorithms for independent assessment of screening mammograms. JAMA Oncol, August 2020.

21. Alejandro Rodriguez-Ruiz, Kristina Lång, Albert Gubern-Merida, Mireille Broeders, Gisella Gennaro, Paola Clauser, Thomas H Helbich, Margarita Chevalier, Tao Tan, Thomas Mertelmeier, Matthew G Wallis, Ingvar Andersson, Sophia Zackrisson, Ritse M Mann, and Ioannis Sechopoulos. Stand-Alone artificial intelligence for breast cancer detection in mammography: Comparison with 101 radiologists. J. Natl. Cancer Inst., 111(9):916–922, September 2019.

22. Michiro Sasaki, Mitsuhiro Tozaki, Alejandro Rodríguez-Ruiz, Daisuke Yotsumoto, Yumi Ichiki, Aiko Terawaki, Shunichi Oosako, Yasuaki Sagara, and Yoshiaki Sagara. Artificial intelligence for breast cancer detection in mammography: experience of use of the Screen-Point medical transpara system in 310 japanese women. Breast Cancer, February 2020.

23. Xiaoqin Wang, Gongbo Liang, Yu Zhang, Hunter Blanton, Zachary Bessinger, and Nathan Jacobs. Inconsistent performance of deep learning models on mammogram classification. J. Am. Coll. Radiol., 17(6):796–803, June 2020.

24. Jeff Donahue, Yangqing Jia, Oriol Vinyals, Judy Hoffman, Ning Zhang, Eric Tzeng, and Trevor Darrell. Decaf: A deep convolutional activation feature for generic visual recognition. In Proceedings of the 31st International Conference on International Conference on Machine Learning - Volume 32, ICML’14, page I–647–I–655. JMLR.org, 2014.

25. A. Gretton, AJ. Smola, J. Huang, M. Schmittfull, KM. Borgwardt, and B. Schölkopf. Covariate shift and local learning by distribution matching, pages 131–160. MIT Press, Cambridge, MA, USA, 2009.

26. A. Torralba and A. A. Efros. Unbiased look at dataset bias. In CVPR 2011, pages 1521– 1528, 2011. doi: 10.1109/CVPR.2011.5995347.

27. Eun-Kyung Kim, Hyo-Eun Kim, Kyunghwa Han, Bong Joo Kang, Yu-Mee Sohn, Ok Hee Woo, and Chan Wha Lee. Applying data-driven imaging biomarker in mammography for breast cancer screening: Preliminary study. Sci. Rep., 8(1):2762, February 2018.

28. B. Efron. Bootstrap methods: Another look at the jackknife. The Annals of Statistics, 7(1):1–26, 1979. ISSN 00905364.

29. PyTorchLightning. PyTorchLightning/pytorch-lightning. https://github.com/PyTorchLightning/pytorch-lightning. Accessed: 2020-6-19.

30. Katie M O’Brien, Stephen R Cole, Chiu-Kit Tse, Charles M Perou, Lisa A Carey, William D Foulkes, Lynn G Dressler, Joseph Geradts, and Robert C Millikan. Intrinsic breast tumor subtypes, race, and long-term survival in the carolina breast cancer study. Clin. Cancer Res., 16(24):6100–6110, December 2010.

31. M Ellingjord-Dale, L Vos, K V Hjerkind, A Hjartåker, H G Russnes, S Tretli, S Hofvind, I Dos-Santos-Silva, and G Ursin. Alcohol, physical activity, smoking, and breast cancer subtypes in a large, nested Case-Control study from the norwegian breast cancer screening program. Cancer Epidemiol. Biomarkers Prev., 26(12), December 2017.

32. Australian Government Department of Health. tPosition statement on breast density and screening within the BreastScreen australia program. September 2016.

33. Sheng Wang, Jiayu Huo, Xi Ouyang, Jifei Che, Xuhua Ren, Zhong Xue, Qian Wang, Jie-Zhi Cheng. mr 2NST: Multi-Resolution and Multi-Reference neural style transfer for mammography. https://arxiv.org/pdf/2005.11926.pdf, May 2020. Accessed: 2020-6-3.

34. Ken Chang, Niranjan Balachandar, Carson Lam, Darvin Yi, James Brown, Andrew Beers, Bruce Rosen, Daniel L Rubin, and Jayashree Kalpathy-Cramer. Distributed deep learning networks among institutions for medical imaging. Journal of the American Medical Informatics Association : JAMIA, 25(8):945–954, 2018. ISSN 1527-974X.

35. Micah J Sheller, G. Anthony Reina, Brandon Edwards, Jason Martin, and Spyridon Bakas. Multi-institutional deep learning modeling without sharing patient data: A feasibility study on brain tumor segmentation. Brainlesion: Glioma, Multiple Sclerosis, Stroke and Traumatic Brain Injuries, 11383:92–104, 2019. ISSN 0302-9743.

## References

1. Welcome to python.org. https://www.python.org/,. Accessed: 2020-6-26.

2. Pydicom |. https://pydicom.github.io/,. Accessed: 2020-6-24.

3. Wu N, Phang J, Park J et al. Deep neural networks improve radiologists’ performance in breast cancer screening. - PubMed - NCBI. https://www.ncbi.nlm.nih.gov/pubmed/31603772. Accessed: 2020-3-6.

4. DICOM standard. https://www.dicomstandard.org/,. Accessed: 2020-6-26.

5. C.11.2 VOI LUT module. http://dicom.nema.org/medical/dicom/current/output/chtml/part03/sect_C.11.2.html,. Accessed: 2020-6-26.

6. nyukat. nyukat/breast_cancer_classifier. https://github.com/nyukat/breast_cancer_classifier. Accessed: 2020-2-9.

7. Histogram matching — skimage v0.17.dev0 docs. https://scikit-image.org/docs/dev/auto_examples/color_exposure/plot_histogram_matching.html,. Accessed: 2019-10-23.

8. William Gale, Luke Oakden-Rayner, Gustavo Carneiro, Andrew P. Bradley, and Lyle J. Palmer. Detecting hip fractures with radiologist-level performance using deep neural networks, 2017.

9. Diederik P Kingma and Jimmy Ba. Adam: A method for stochastic optimization. December 2014.

10. NVIDIA. NVIDIA/apex. https://github.com/NVIDIA/apex. Accessed: 2020-6-23.

11. Sage Bionetworks. Synapse | sage bionetworks. https://www.synapse.org/#!Synapse:syn9773040/wiki/426908. Accessed: 2020-6-23.

